# Influenza season 2019: analysis of 143 hospitalized cases

**DOI:** 10.1101/2020.09.16.20195974

**Authors:** Indalecio Carboni Bisso, Eduardo Prado, Joaquin Cantos, Agustín Massó, Inés Staneloni, Eduardo San Román, Iván Huespe, Marcos Las Heras

**Affiliations:** Intensive Care Unit, Hospital Italiano de Buenos Aires; Infectious Diseases, Hospital Italiano de Buenos Aires

## Abstract

**Introduction:** Influenza virus infection is a latent public health problem, affecting millions of people through the planet, and it is an important cause of morbidity and mortality. In Argentina, there is a significant absence of data regarding influenza severe respiratory disease and, therefore, a lack of knowledge about the impact of this disease at health institutions.

**Objectives:** Analysis of clinical characteristics, image findings and laboratory variables in patients with influenza viruses during 2019.

**Methods:** Retrospective, single-centre study, we analyzed all confirmed cases of influenza in a high complexity hospital from Buenos Aires.

**Results:** 143 patients with influenza virus were hospitalized in this period of time. The 98.6% were infected by type A influenza, and most of them 61.5% were H1N1 subtype.

Median age was 71 years (IQR 60 - 82), 77.6% were older than 70 years, and 88.1% had at least one coexisting illness. 39.1% of the patients required intensive care, 11.1% invasive mechanical ventilation and 4.1% died during hospitalization.

**Conclusion:** Mortality and severity were similar to previous series of non-pandemic influenza. Analysis of annual data would be valuable in order to document the severity of influenza hospitalizations by ageJgroup and comorbidities according to the circulating influenza viruses.

## INTRODUCTION

Influenza virus infection is a latent public health problem, affecting millions of people through the planet, and it is an important cause of morbidity and mortality, especially for certain susceptible populations.

Influenza virus is a single stranded RNA virus, characterized by 4 types (A, B, C and D). This virus has 2 important surface glycoproteins, hemagglutinin (H1, H2 and H3, the most common in humans) and neuraminidase (N1 and N2, the most common in humans), which are used to classify the different subtypes and give the virus a changing antigenicity as its main characteristic. The term “antigenic drift” makes reference to the periodic changes in the antigenicity of the virus that occur year by year, while “antigenic shift” describes the generation of large genetic recombinations which are responsible for pandemics, such as the one that took place in 2009 ^1^.

Influenza infection can evolve, due to the same pathogenicity of the virus or bacterial superinfection, into a severe respiratory infection. Among the risk factors for presenting a severe influenza disease, it is possible to recognise increasing age, cardiovascular disease, malignancy, immunosuppression, hypertension, neuromuscular disease, diabetes mellitus, chronic lung, liver, or metabolic renal disease. Also obesity and pregnancy were associated with increased risk of negative outcomes in some studies ^2,3^.

Among the patients hospitalized due to influenza severe respiratory infection, it has been estimated that 29 to 6% require admission to the intensive care unit (ICU). A recent multicentre European study which analyzed the characteristics and outcomes of hospitalized patients with diagnosis of influenza from 2009 to 2017 reported that 43% of the patients admitted to the ICU did not have predisposing factors and 21% of hospital mortality.

In Argentina, there is a significant absence of data regarding influenza severe respiratory disease and, therefore, a lack of knowledge about the impact of this disease at health institutions, hospital mortality, and the profile of patients requiring ICU. Thus, the objective of this work is to describe the history of comorbidities as well as the clinical, laboratory and imaging findings of patients who required hospitalization in a general ward or ICU during 2019 in a high-complexity care hospital from Buenos Aires, capital of Argentina.

## METHODS

In this retrospective, observational single-centre study, we included patients from January 1 to December 31 2019 at a high complexity hospital from Buenos Aires. The study was approved by the Ethics Committee from our hospital in October 2019.

### Data sources

We obtained epidemiological, demographic, clinical, laboratory, management, and outcome data from patients’ medical records. Clinical outcomes were followed up to march, 2020.

### Procedures

Throat-swab specimens from the upper respiratory tract that were obtained from all patients at admission were maintained in viral-transport medium. Influenza and other respiratory viruses including adenovirus, respiratory syncytial virus, parainfluenza viruses, rhinovirus, enterovirus, coronavirus and bocavirus were confirmed by using real time reverse transcription polymerase chain reaction (real time RT-PCR). Due to sequence similarity, rhinovirus and enterovirus may not be discriminated by the technique used. Sputum or endotracheal aspirates were obtained at admission for identification of possible causative bacteria or fungi. Additionally, all patients were given chest x-rays or chest computed tomography scans (CTs).

### Outcomes

We describe epidemiological data, demographics, signs and symptoms on admission, comorbidity, laboratory results; co-infection with other respiratory pathogens, chest radiography and CT findings, treatment received and clinical outcomes.

### Statistical analysis

Continuous variables were expressed as medians and interquartile ranges or simple ranges, as appropriate. Categorical variables were summarized as counts and percentages. No imputation was made for missing data. Because the cohort of patients in our study was not derived from random selection, all statistics are deemed to be descriptive only. We used RStudio developed by R-Tools Technology Inc for all analyses.

## RESULTS

143 patients with influenza were included in this study, 79 patients were female and 64 men. Median age was 71 years (IQR 60 - 82). Among the overall population, 77.6% (111) were older than 70 years, and 88.1% (126) had at least one coexisting illness. Chronic obstructive pulmonary disease (COPD) was the most frequently observed (30.0%), followed by immunosuppression (27.2%) and cardiovascular disease (19.5%). 56 patients (39.9%) required ICU admission and the rest of them were treated in general ward. 16 patients (11.1%) underwent invasive mechanical ventilation (iMV), 40 (27.9%) non invasive ventilation or high-flow nasal cannula and 6 patients (4.1%) died during hospitalization. Furthermore, 5 patients (9%) hospitalized in ICU died.

Vaccination status was known in all cases, with a vaccination rate of 32.1% (28) in non-ICU patients and 26.7% (15) in ICU patients. 6.9% (3) of vaccinated patients required iMV vs. 13% (13) of the unvaccinated cases. The median hospitalization time was 7 days (IQR 4 - 13) for all patients, 5 days (IQR 3 - 10) for non-ICU patients and 10 days (IQR 6 - 16) for ICU cases. Demographic and clinical characteristics of patients are shown in *Table 1*.

**Table 1.**
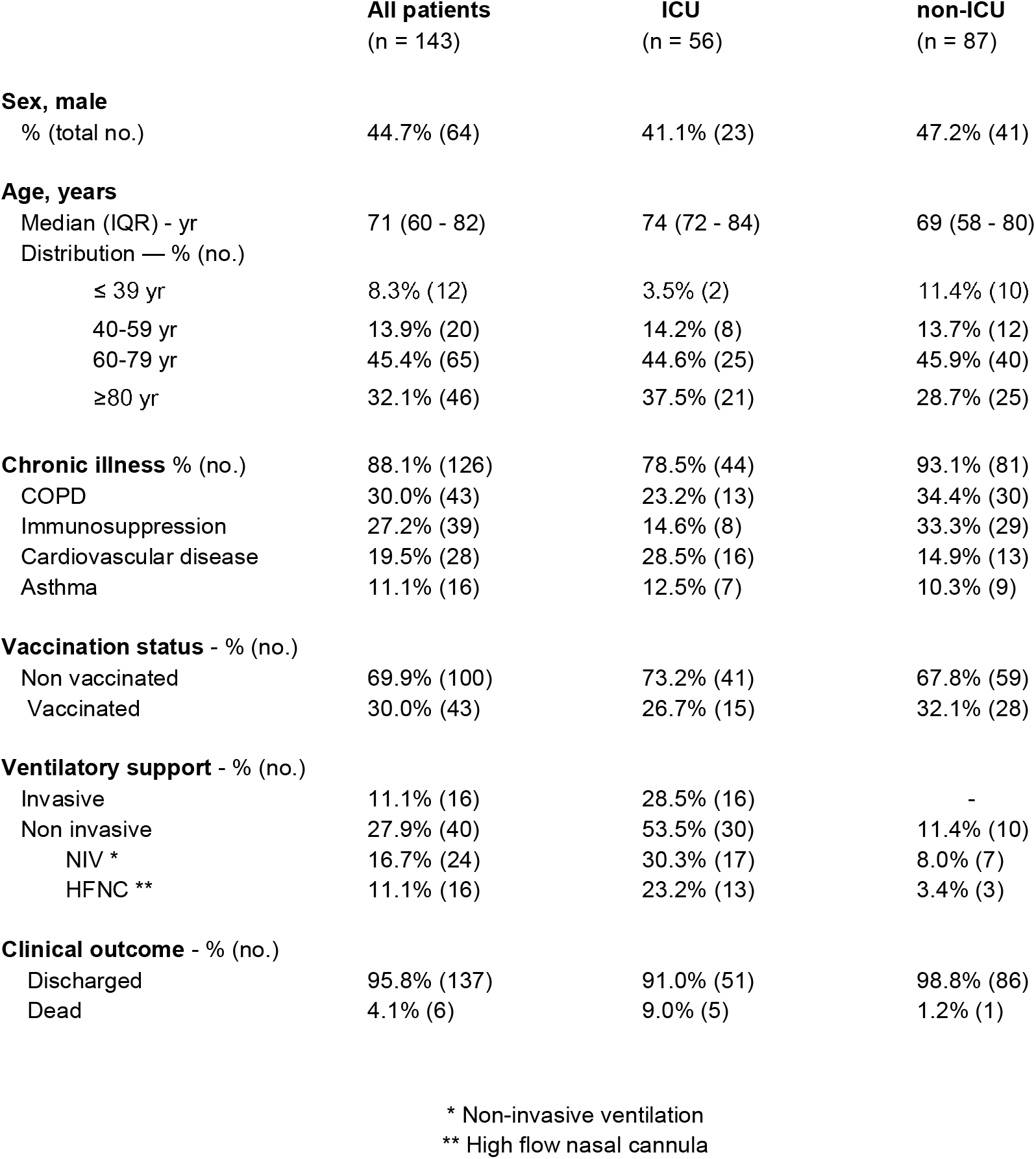
Patients characteristics.

|  | All patients<br>(n = 143) | ICU<br>(n = 56) | non-ICU<br>(n = 87) |
| --- | --- | --- | --- |
| **Sex, male** |  |  |  |
| % (total no.) | 44.7% (64) | 41.1% (23) | 47.2% (41) |
| **Age, years** |  |  |  |
| Median (IQR) - yr | 71 (60 - 82) | 74 (72 - 84) | 69 (58 - 80) |
| Distribution — % (no.) |  |  |  |
| ≤ 39 yr | 8.3% (12) | 3.5% (2) | 11.4% (10) |
| 40-59 yr | 13.9% (20) | 14.2% (8) | 13.7% (12) |
| 60-79 yr | 45.4% (65) | 44.6% (25) | 45.9% (40) |
| ≥80 yr | 32.1% (46) | 37.5% (21) | 28.7% (25) |
| **Chronic illness % (no.)** | 88.1% (126) | 78.5% (44) | 93.1% (81) |
| COPD | 30.0% (43) | 23.2% (13) | 34.4% (30) |
| Immunosuppression | 27.2% (39) | 14.6% (8) | 33.3% (29) |
| Cardiovascular disease | 19.5% (28) | 28.5% (16) | 14.9% (13) |
| Asthma | 11.1% (16) | 12.5% (7) | 10.3% (9) |
| **Vaccination status - % (no.)** |  |  |  |
| Non vaccinated | 69.9% (100) | 73.2% (41) | 67.8% (59) |
| Vaccinated | 30.0% (43) | 26.7% (15) | 32.1% (28) |
| **Ventilatory support - % (no.)** |  |  |  |
| Invasive | 11.1% (16) | 28.5% (16) | - |
| Non invasive | 27.9% (40) | 53.5% (30) | 11.4% (10) |
| NIV * | 16.7% (24) | 30.3% (17) | 8.0% (7) |
| HFNC ** | 11.1% (16) | 23.2% (13) | 3.4% (3) |
| **Clinical outcome - % (no.)** |  |  |  |
| Discharged | 95.8% (137) | 91.0% (51) | 98.8% (86) |
| Dead | 4.1% (6) | 9.0% (5) | 1.2% (1) |

\* Non-invasive ventilation
\*\* High flow nasal cannula

Influenza viruses were detected by real-time RT-PCR. 98.6% of patients were infected by type A influenza, 61.5% had subtype H1N1 and 37.0% H3N2. Most of the cases were reported during the 24th to 32nd epidemiological weeks of winter season in the Southern hemisphere. Annual distribution of the cases is detailed in *Figure 1* and the distribution of cases by viral type and subtype in *Figure 2*.

**Figure 1.**
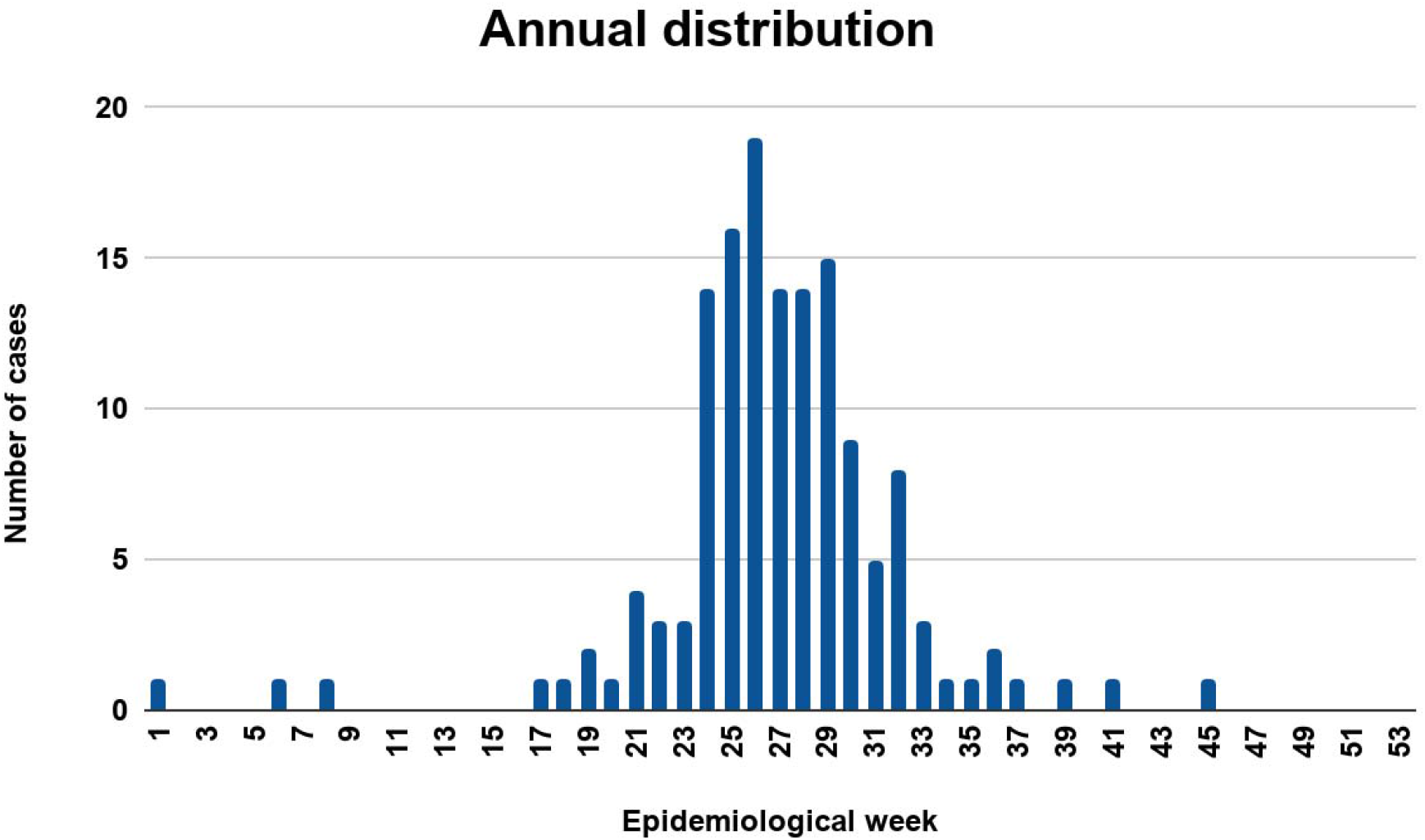
Annual distribution of influenza cases.

**Figure 2.**
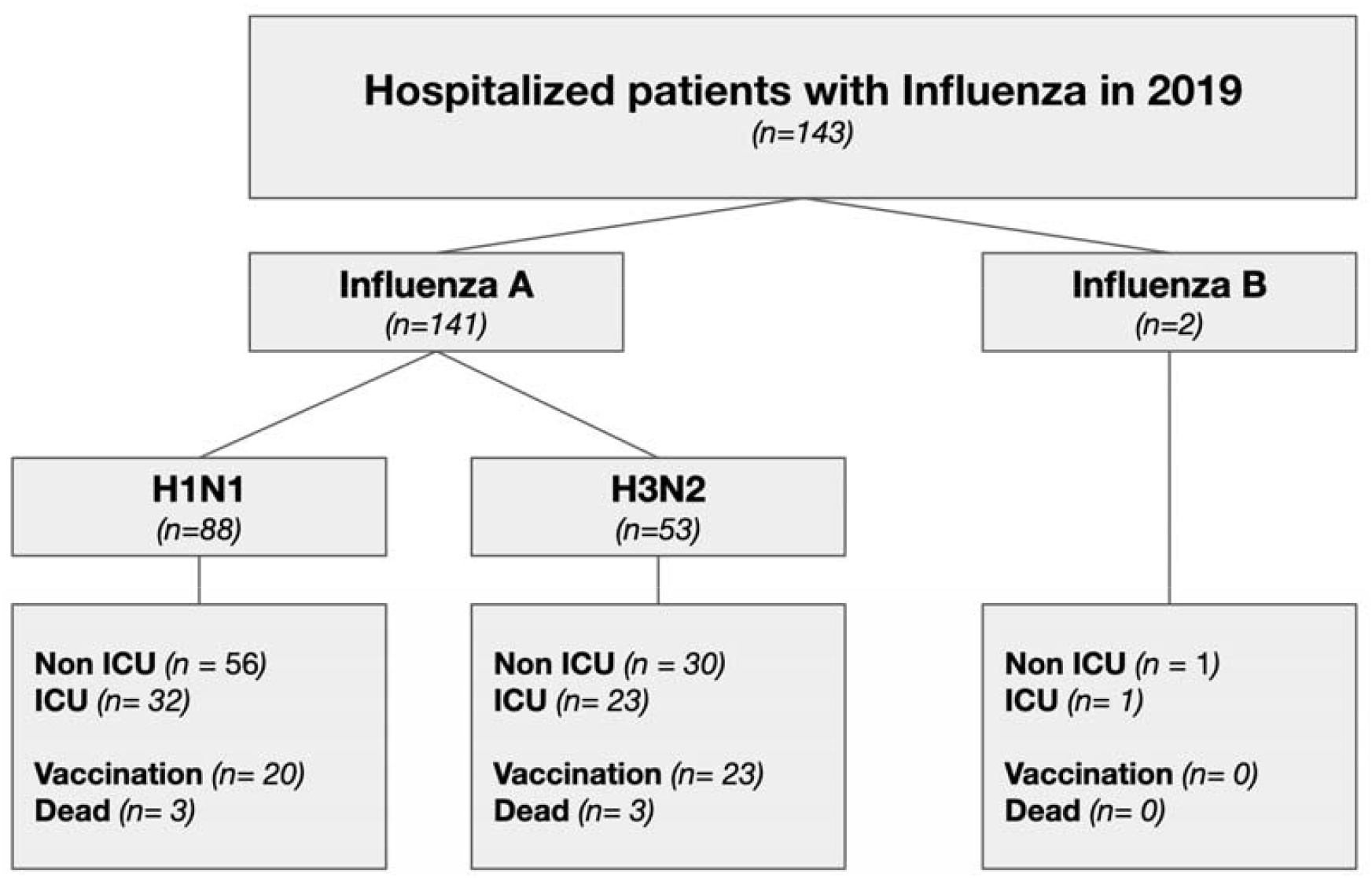
Distribution of cases by viral type and subtype.

Regarding clinical findings, fever was present on admission in 65.0% (93) of the patients. The second most common symptom was shortness of breath (58.0%), followed by cough (52.4%) and rhinorrhoea (19.5%) diarrhoea, nausea or vomiting (4.8%) were uncommon. Also dyspnoea was more frequent in ICU patients (69.6% vs. 50.5%).

Among laboratory findings, lymphocytopenia was present in 64.3% of the patients, leukocytosis in 35.6% and neutrophilia in 24.4%. Hyponatremia was found in 46.1% of the patients, and 28.6% presented elevated serum creatinine and urea. Half of the patients [50.0% (28)] who underwent admission to ICU presented elevated serum pro b-type natriuretic peptide (proBNP) (median 2044 pg/ml, IQR 781 - 4904) that was also elevated in 12.6% (11) of the non-ICU patients. In addition, elevation of serum transaminases was more frequently seen in ICU patients (37,5% vs. 8.0%), without associated involvement of alkaline phosphatase, bilirubin or prothrombin time. *Table 2* shows clinical presentation and laboratory findings.

**Table 2.**
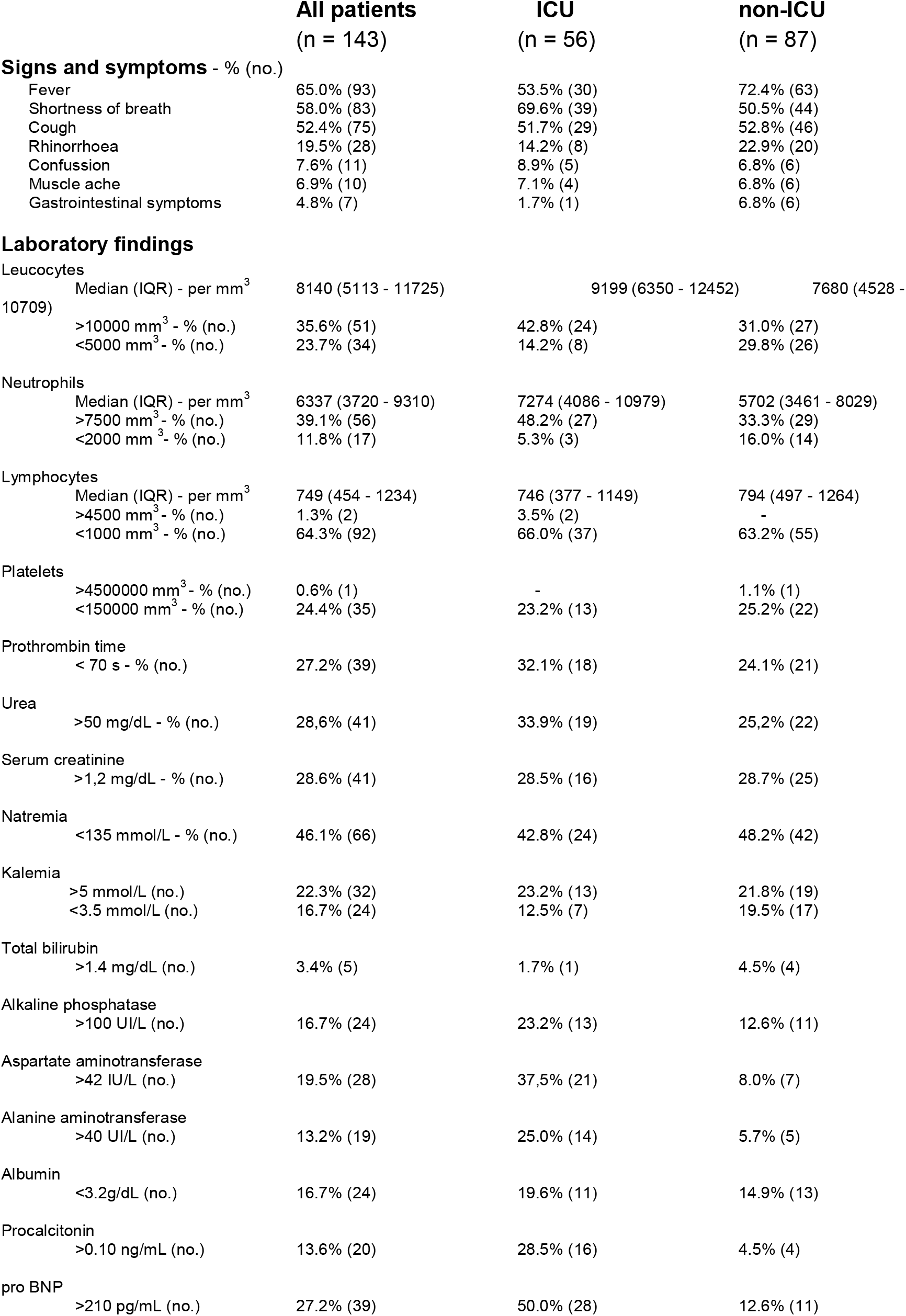
Clinical presentation and laboratory findings.

Of 129 chest x-rays that were performed at the time of admission, 67.4% revealed abnormal results. The most common patterns shown were: interstitial opacity (35.6%), consolidation (24.8%) and pleural effusion (6.9%). Also, 33.3% of the patients presented bilateral compromise. Moreover, almost the total of ICU cases [98.1% (50)] showed pathological findings in chest x-rays in contrast with admissions to general ward [47.4% (37)].

85 chest CTs were performed. No CTs abnormality was found in 11 (12.9%) studies. Ground-glass opacity was found in 34.1%, followed by consolidation in 29.4%, tree in bud sign in 15.2%, and multiple mottling opacity in 8.2%. In addition, bilateral compromise was informed in 44.7% and pleural effusion in 14.1%. Furthermore, bilateral compromise was more frequent among ICU patients compared with non-ICU patients in both chest x-rays [50.9% (26) vs. 21.7% (17)] and chest CTs [67.6% (23) vs. 29.4% (15)]. *Table 3* shows images findings.

**Table 3.**
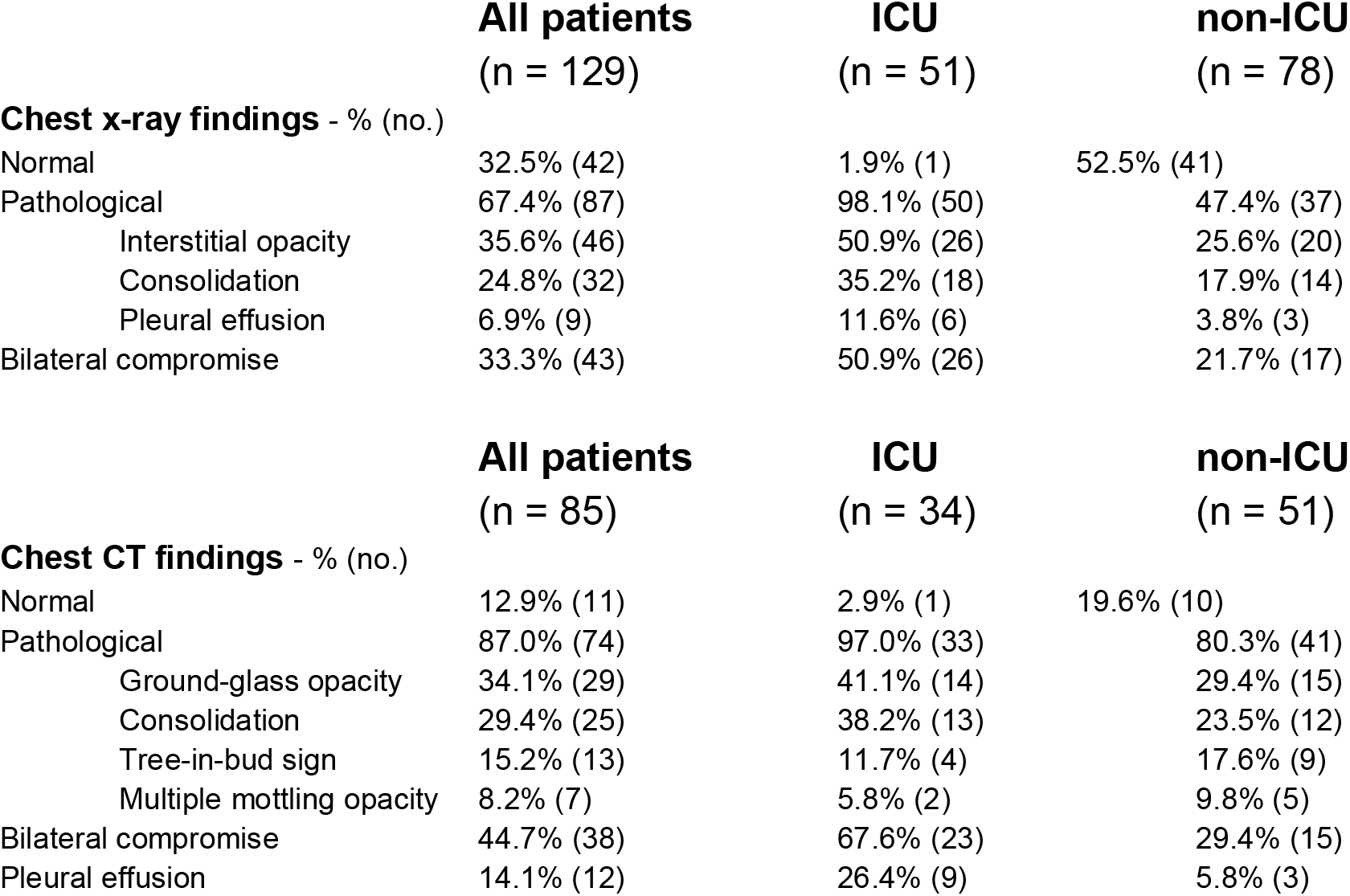
Images findings.

## DISCUSSION

In this cohort study, we reported the clinical characteristics and risk factors associated with clinical outcomes in patients with laboratory-confirmed influenza who required hospitalization during 2019. We found that overall mortality was similar to previous series. However, in ICU cases, mortality was significantly lower (9% vs. 10 to 31% range), despite the fact that the majority of the cases reported in this study were older than 70 years and had at least one coexisting illness ^4–6^. Also, the relative amount of patients admitted to ICU was higher. This may be attributed to the ICU criteria of our unit, where age is not a restriction for invasive vital support. Nevertheless, it is important to point out the lack of post-ICU follow up, therefore, we do not evaluate cognitive, emotional or physical status after discharge.

In this series, vaccination acted as a protective factor against severe illness, with less ICU admission, less iMV requirement and a decreased length of stay. It is well documented that influenza vaccines provide substantial protection against severe influenza illness among adults. But also, they seem to provide low protection for elderly patients in seasons where vaccine and circulating A(H3N2) strains were antigenically variant ^7,8^. During 2019, a double viral circulation was registered in Argentina with the presence of subtypes H1N1 and H3N2. These findings should be taken into account to improve vaccination strategies and achieve better vaccination coverage in order not only to decrease flu cases, but also their severity.

As regards chronic medical illnesses, we found a lower quantity of immunosuppressed patients in ICU against the group that required hospitalization in general ward (14.6% vs. 33.3%). Additionally, mortality was higher among immunocompetent patients (3.8% vs. 0.5%) with no difference in median length of stay or iMV requirement. In this sense it is reported that a lower inflammatory response in immunosuppressed patients against the pathogen is associated with better outcomes and less organic failure ^9,10^. It is interesting to analyze the behavior of influenza in immunosuppressed patients in order to draw conclusions and extrapolate information to the general of this population. However, we believe that it is worth highlighting these findings that we mentioned in order to inform professionals and undertake future research.

In relation to the laboratory findings, hepatic enzymes impairment, mainly ASAT, was far more frequent in ICU cases than in general patients. In a recently published research, ASAT rate superior to 68 IU/L was identified as a predictor of 3-month mortality with other variables such as creatinine and PaO2/FiO2 with a hazard ratio (HZR) of 7.68 (IC 95% 1.6-35.1), the highest HZR among them ^11^. Additionally, proBNP was higher in ICU patients (median 2044 pg/mL, IQR 781 - 4904 vs. 1361, IQR 537 - 2341), in fact 50.0% of critical patients had values higher than normal range (210 pg/mL) vs. 12.6% of non ICU patients. ProBNP has been reported to be useful as a predictor in patients with confirmed influenza ^12^.

Regarding images studies, it is important to pinpoint that in our series only one patient of those who required critical care had a normal result. Likewise, greater radiological impact was seen in ICU cases, with more bilateral compromise in both chest x-rays (50.9% vs. 21.7%) and chest CTs (26.4% vs. 5.8%) in comparison with non-ICU patients. Similar data was found in other series where a high percentage of pathological radiographic findings were detected in patients with a worse prognosis ^13–15^.

Our study has several limitations. It is an observational study of a single center and data from other Argentine centers are necessary to know the impact of the influenza in hospitalizations. Also, as previously noted, no post-ICU follow up was performed.

Regarding the behavior of the virus in 2019, we conclude that both mortality and severity of the cases were similar to those published in previous series of non-pandemic influenza. Analysis of annual data would be valuable in order to document the severity of influenza hospitalizations by age group and comorbidities according to the circulating influenza viruses.

## Data Availability

All relevant data are within the manuscript and its Supporting Information files.

